# Road networks to explore COVID-19 infection

**DOI:** 10.1101/2023.01.31.23285228

**Authors:** Shahadat Uddin, Arif Khan, Haohui Lu, Fangyu Zhou, Shakir Karim, Farshid Hajati, Mohammad Ali Moni

**Affiliations:** School of Project Management, Faculty of Engineering, The University of Sydney, Forest Lodge, NSW 2037, Australia; College of Engineering and Science, Victoria University Sydney, Sydney, NSW 2000, Australia; School of Health and Rehabilitation Sciences, Faculty of Health and Behavioural Sciences, The University of Queensland, St Lucia, QLD 4072

**Keywords:** Road networks, Infection count, Socio-economic factors, COVID-19

## Abstract

COVID-19 pandemic triggered an unprecedented level of restrictive measures globally. Most countries resorted to lockdowns at some point to buy the much-needed time for flattening the curve and scaling up vaccination and treatment capacity. Although lockdowns, social distancing and business closures generally slowed down the case growth, there is a growing concern about the social, economic and psychological impact of these restrictions, especially on the disadvantaged and poorer part of society. While we are all in this together, these segments are often taking the heavier toll of the pandemic and facing harsher restrictions or getting blamed for community transmission. This study tries to explore this perspective using quantitative analysis and network theory. The research is set in the context of the latest delta and omicron outbreaks in the Greater Sydney area, Australia, during late 2021. We first try to model how the local road networks between the neighbouring suburbs (i.e., neighbourhood measure) and current infection count affect the case growth and how they differ between delta and omicron variants. We use a geographic information system, population and infection data to measure - road connections, mobility and transmission probability across the suburbs. We then looked at three socio-demographic variables – age, education and income and explored how they moderate independent and dependent variables (infection rates and neighbourhood measures). The result shows strong model performance to predict infection rate based on neighbourhood road connection. However, apart from age in the delta variant’s context, the other two variables – income and education level do not seem to moderate the relation between infection rate and neighbourhood measure. The results indicate that suburbs with a more socio- economically disadvantaged population do not necessarily contribute to more community transmission. The study findings could be potentially helpful for stakeholders in tailoring any health decision for future pandemics.

## 1. Introduction

COVID-19 pandemic has caused a significant amount of mortality, illnesses and hospitalization as well as impacted transport, logistics (Nižetić 2020) and economies (Štifanić et al., 2020) globally. The pandemic is estimated to cause the greatest recession since the 1930s (the Great Depression) and will possibly cause 420-580 million more people to live in poverty (Sumner et al., 2020). A significant amount of effort has been made in preventing and treating this disease, curbing the growth through various restrictions and public health measures. Many governments invest funds to combat this disease (Haug et al., 2020) and minimize economic losses due to lockdowns and business closure. As for academics, a global effort has been put forward to understand the patho-physical properties of the virus, evaluate public health measures, and model the transmission that could help predict the spread based on historical data.

Many classical models have been implemented using this disease’s spread data, and they have mostly turned out effective in capturing the future trend. Hernandez-Matamoros (2020) evaluated the autoregressive integrated moving average (ARIMA) model with data from 145 countries within six regions and showed its effectiveness for predicting COVID-19 as well. Their paper outlined a relationship between the COVID-19 spread pattern and the population in a region, which showed its potential to build models to predict the COVID-19 transmission using variables such as culture, climate, humidity, etc. Swaraj (2021) proposed an ARIMA-based model that could capture the data’s linear and non-linear components by integrating an autoregressive neural network. The hybrid method exhibited a significant reduction in terms of different performance measures (e.g., root-mean-squared error and mean absolute error) compared to the single ARIMA model for observed cases daily. Some variations of the classical SIR (Susceptible-Infected-Recovery) model were also used. For example, Abdy (2021) proposed a new SIR model with fuzzy parameters like infection rate, recovery rate, and death rate due to COVID-19. Liu (2021) extended the current susceptible-exposed-infected-recovery (SEIR) model, which is a variation of the SIR model, by incorporating extra compartments. This model can explain the new features of COVID-19 and fine-tune the new model with a neural network aimed at a higher accuracy prediction.

As many of the classical statistical models might show their inability to use some unique determining parameters, machine learning models have provided an alternative when understanding much more complicated datasets. Many models have been applied to different datasets on COVID-19, with artificial neural network (ANN) and recurrent neural network (RNN) being the most promising techniques so far. Car et al. (2020) proposed the first ANN-based model to predict the COVID-19 spread trend. They trained three distinct models using confirmed, recovered and deceased cases and achieved 0.94 for the coefficient of determination. Melin et al. (2020) presented a multiple ensemble ANN model where a fuzzy response aggregation for time series data was used. The ensemble ANN models make it possible to predict for various conditions, and a fuzzy logic could help aggregate the responses of these neural predictors. On top of these, the best determination coefficient achieved so far is from the experiments by Pinter et al. (2020), who used ANFIS and MLP-ICA methods to predict the number of infected people and the mortality rates. Their determination coefficient score reached 0.99 when applying the MLP-ICA method. The typical modelling using RNN and the best results among RNN variants are developed from the long short-term memory method (LSTM). Chimmula and Zhang (2020) used an LSTM-based approach to forecast COVID-19 patterns and concluded that the pandemic would come to an end by the end of June 2020. Such a conclusion could be considered quite plausible only for the COVID-19 first wave. Yudistira (2020) also used LSTM to understand and model the correlation of the COVID-19 growth rate. The optimal structure of the models was determined heuristically. Their experiments concluded that LSTM outperformed RNN when using RMSE value as the comparing metrics.

As we know, most governments employ some sort of restrictions on people’s mobility to protect public health (Varotsos and Krapivin 2020). These regulations varied considerably in terms of guidelines, duration and geographical coverage based on various economic, social and public health factors. Although such restrictions have been used during earlier epidemics in various times and places, the current COVID-19 pandemic is notably different from similar historical events. COVID-19 has high transmissibility and frequently mutates (Lotfi et al., 2020). There has been a limited study to verify to what extent these restrictions on mobility and business closures are providing in terms of cost and benefit, and also whether there could be other factors (e.g., income level, economic support, awareness, education etc.), if improved, could be more effective than mobility restriction in order to fight the virus.

This study will be utilizing a network-based approach and panel regression methods to analyze the effects of human mobility in transmitting COVID-19, together with a close look at a suburban population’s characteristics like their age, income and education. Notably, the mobility in this work will be represented by the actual roads between suburban areas, given the assumptions that more roads would usually result in a higher level of mobility.

## 2. Our approach

As summarised in the Introduction section, researchers used a wide range of attributes to model the number of COVID-19 infections for a geographic area in a given period. Due to the highly infectious nature of COVID-19, this study considered features that affect the direct transmission of the virus between individuals. There is a good chance of a higher number of future COVID-19 infections in a suburb if it already has an increased number of infected residents. Similarly, the possibility for the same suburb to have a higher number of infected patients will increase if it has direct road connections with suburbs with many COVID-19 infected patients. It would be difficult to control human mobility entirely at the inter suburban level; even strict lockdown or curfew will be in place (Zhou et al., 2020, Al Wahaibi et al., 2021).

Accordingly, this study considered two time-series measures to model the COVID-19 infection number for a given postal area or suburb. The first one is the infection number or count from previous time points. The second one is a composite one and is based on the suburban road network. It is a weighted sum based on the number of road connections to each neighbouring suburb (i.e., the weighting factor) and their respective infection count at the previous time point. The following formula can capture our approach.

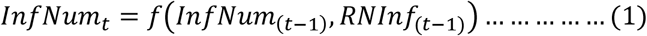

Where *InfNum*_*t*_ is the number of infected COVID-19 patients in a suburb at time *t* (i.e., current infection number), *InfNum*_(t-1 ;)_ is the number of infected COVID-19 patients at time *(t-1)* (i.e., previous infection number), and *RNInf*_(t-1)_ is the road network-based infection measure at *(t-1)* (i.e., neighbourhood measure). Mathematically, the following formula represents this measure.

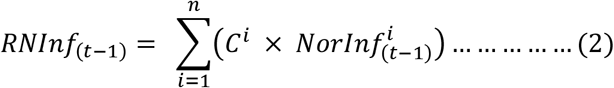

Where *n* indicates the number of other suburbs that the underlying suburb has road connections, *C*^*i*^ is the number of road connections the suburb has with the suburb *i*, and 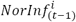 is the normalized infection number of suburb *i* at (t − 1) time point. This study considers the population sizes of the neighbouring suburbs to normalize their respective infection numbers. Since this measure depends on its connection with neighbouring suburbs and their infection number for a given suburb, this study names it the *neighbourhood measure*.

## 3. Methods and materials

### 3.1 Data source

This study considered the COVID-19 infection data for 100 different suburbs of the Greater Sydney area of New South Wales, Australia (NSW Health 2021). We considered two distinct periods for the infection statistics of these suburbs: one for the delta variant (four weeks starting from August 24, 2021) and another for the omicron variant (four weeks beginning on November 17, 2021). The delta variant also spread during the second period. However, we termed this period as ’omicron’ since the omicron variant had already become prevalent in these suburbs from early November 2021 (NSW Health 2021). Table 1 details the basic statistics of the infection data considered in this study.

**Table 1:** The basic statistics of the COVID-19 infection data for 100 suburbs considered in this study.

| **Omicron** | Overall | Week 1<br>(17-23 Nov 2021) | Week 2<br>(24-30 Nov 2021) | Week 3<br>(1-7 Dec 2021) | Week 4<br>(8-15 Dec 2021) |
| --- | --- | --- | --- | --- | --- |
| Mean | 8.36 | 2.80 | 4.57 | 6.81 | 19.27 |
| Change (%) | - | - | 63% | 49% | 183% |
| Standard Deviation | 13.72 | 3.96 | 6.66 | 9.85 | 20.81 |
| Sample Variance | 188.21 | 15.68 | 44.41 | 97.04 | 433.03 |
| Minimum | 0 | 0 | 0 | 0 | 0 |
| Maximum | 104 | 24 | 43 | 65 | 104 |

| **Delta** | Overall | Week 1<br>(24-30 Aug 2021) | Week 2<br>(31 Aug – 6 Sep 2021) | Week 3<br>(7-13 Sep 2021) | Week 4<br>(14-20 Sep 2021) |
| --- | --- | --- | --- | --- | --- |
| Mean | 49.69 | 63.36 | 55.43 | 45.15 | 36.48 |
| Change (%) | - | - | -13% | -19% | -19% |
| Standard Deviation | 63.54 | 77.15 | 65.78 | 58.87 | 46.89 |
| Sample Variance | 4037.19 | 5951.93 | 4327.12 | 3465.48 | 2198.80 |
| Minimum | 0 | 0 | 0 | 0 | 0 |
| Maximum | 439 | 439 | 372 | 347 | 239 |

To quantify the second independent variable (*RNInf*_(t-1)_), we first construct the suburban road network. A node in this network represents a suburb. An edge between two nodes indicates at least one road connecting the underlying suburbs represented by those nodes, and the edge weight points to the number of roads connecting the two suburbs of the edge. We took the map data from Google Maps, Australia (Google maps 2021). Figure 1 illustrates an example of the suburban road network construction. For a given suburb, we then considered the infection number for each of its neighbouring suburbs. Finally, we used formula (2) to quantify this measure.

**Figure 1:**
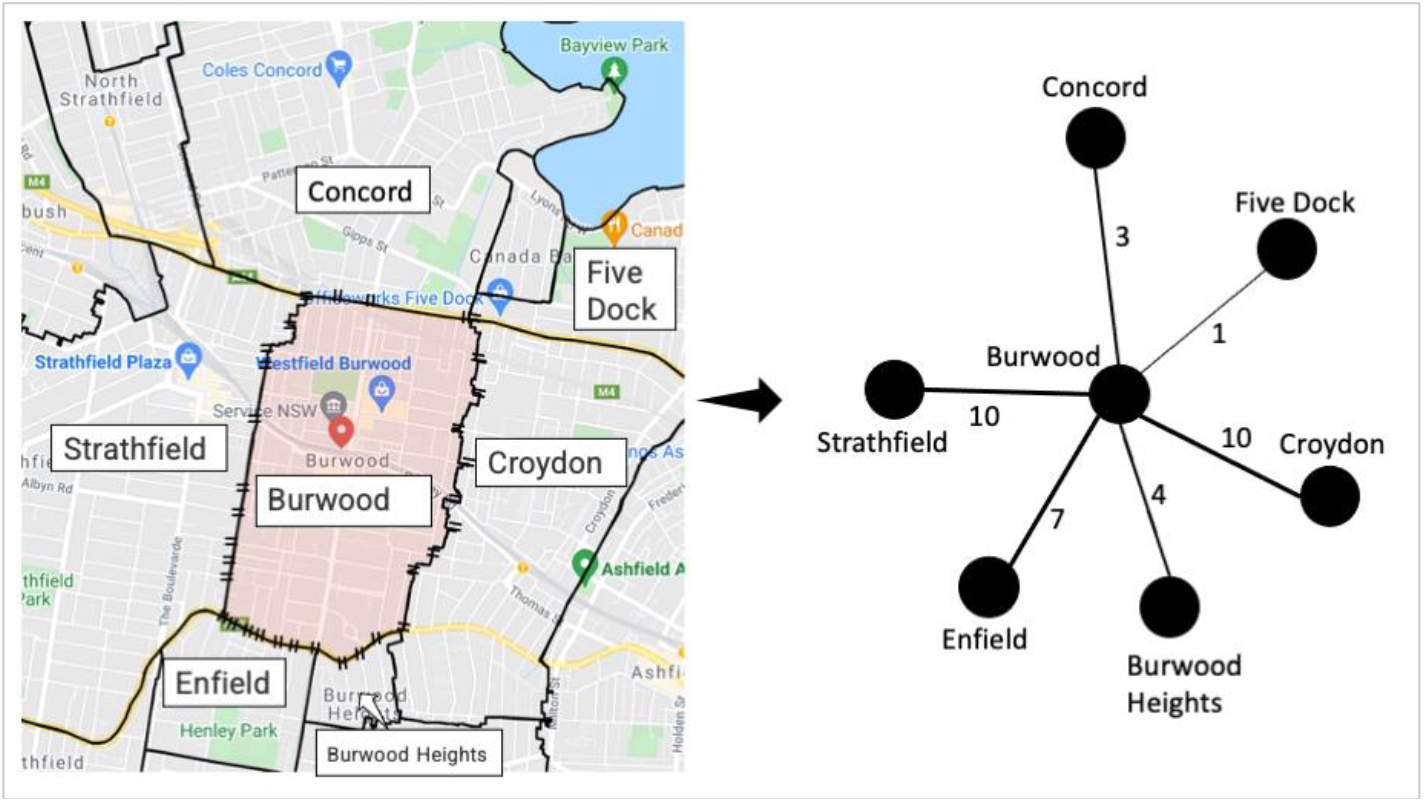
An illustration of the construction of the suburban road network. The left-hand figure shows the map from the Google Maps website. The right-hand figure is the corresponding suburban road network. Burwood (shaded with light red colour) is the suburb under consideration. Edge weights between two suburbs are the number of roads connecting them. For example, the edge weight (right-hand figure) between Burwood and Strathfield is ten since ten roads connect these two suburbs (left-hand figure). Edge thickness in the right-hand figure proportionates to the corresponding edge weight.

This study considered three moderating attributes (i.e., age, education and income) to investigate their impact on the relationship between the dependent and independent variables of this study’s proposed model. The relevant data of these two socio-demographic attributes for different suburbs were collected from the census data provided by the Australian Bureau of Statistics (Census QuickStats 2021).

### 3.2 Data analysis design

Since this study repeatedly measured the model’s variables four times, we followed the panel regression to explore the proposed model. We considered one week for each repeated measure. In particular, we used fixed effect panel regression for research data analysis since we found a significant correlation between the error terms and the independent variables from the initial data exploration. We used Stata to run the fixed effect panel regression (Kohler and Kreuter 2005).

This study considered the median population age value, the percentage of residents having a university or tertiary degree, and the median weekly household income to measure the three socio-demographic attributes, age, education and income, respectively, for each suburb. The median values for *age, education* and *income* attributes for 100 data instances have split the dataset into two groups. For example, the *education=0* group includes all suburbs with a lower percentage of residents having a university degree than the median value of all data instances of this study, and vice versa. We first created six more independent variables to check their moderating strength by multiplying each with the first two independent variables (i.e., InfNum_(t-1)_ and RNInf_(t-1)_). Then, we reran the panel regressions, including these six newly created independent variables.

## 4. Results

Figure 2 illustrates the undirected road network among the 100 suburbs considered in this study. In this network, there are 214 undirected edges among its 100 nodes. The maximum number of roads connecting two suburbs is 16, between 2142 and 2160 postal areas.

**Figure 2:**
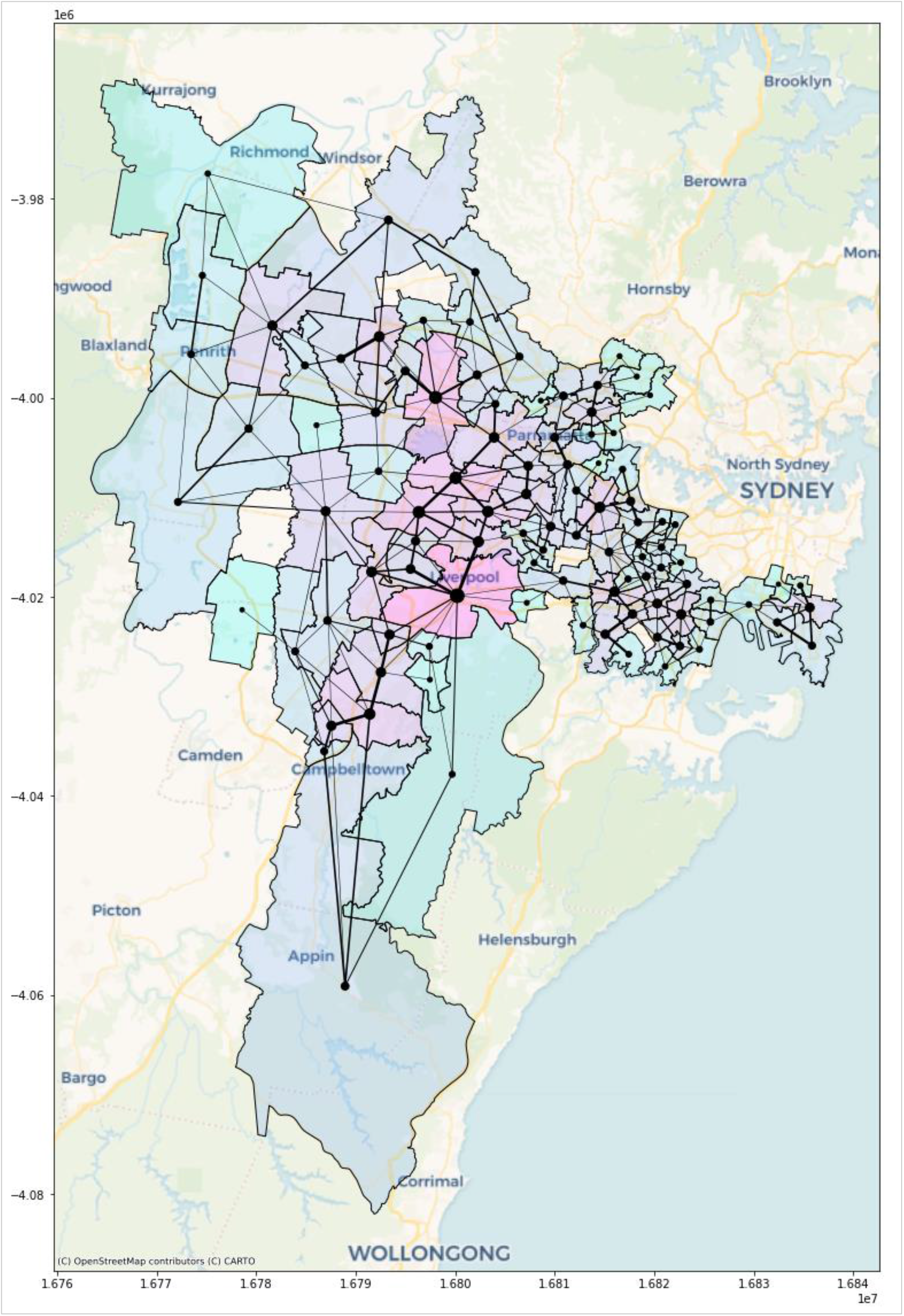
The road network among the 100 suburbs considered in this study. The node’s size proportionates to its degree of centrality (i.e., the number of connections it has with its neighbouring suburbs) in the network. The edge thickness between two nodes is proportional to the number of roads connecting the corresponding suburbs represented by those two nodes (Map projection: Web Mercator)

Table 2 shows the results from the fixed effect panel regressions. The models for both omicron and delta variants show very high R-squared values. The R-squared value for the delta variant is 0.8566, and for the omicron variant, it is 0.5267. Previous infection number (InfNum_(t-1)_) significantly impacts the present infection number for the delta and omicron variants. Neighbourhood measure (RNInf_(t-1)_) also significantly impacts the present infection number. It shows a positive impact on the delta variant. However, it shows a negative impact on the omicron variant.

**Table 2:** Panel regression outcome for delta and omicron variants

|  | Delta |  |  |  | Omicron |  |  |  |
| --- | --- | --- | --- | --- | --- | --- | --- | --- |
| Independent variable | Coef. | Std. Err. | t-Statistic | Sig. | Coef. | Std. Err. | t-Statistic | Sig. |
| Constant | 8.7439 | 3.2626 | 2.68 | 0.008 | 6.2191 | 1.6186 | 3.84 | 0.000 |
| $InfNum_{(t-1)}$ | 0.5466 | 0.0586 | 9.33 | 0.000 | 1.4319 | 0.1201 | 11.92 | 0.000 |
| $RNInf_{(t-1)}$ | 0.2642 | 0.0909 | 2.91 | 0.004 | -0.1016 | 0.0369 | -2.76 | 0.006 |
| Model parameter |  |  |  |  |  |  |  |  |
| R-squared | 0.8566 |  |  |  | 0.5267 |  |  |  |
| F-statistic | 96.56 |  |  |  | 77.38 |  |  |  |
| Prob (F-statistic) | 0.000 |  |  |  | 0.000 |  |  |  |

To check the moderating impact of three socio-demographic attributes (i.e., age, education and income) on the findings of Table 2, we added six more independent variables to our dataset and repeated the same panel regression. These six composite variables are based on the multiplication of each socio-demographic attribute with the three independent variables. The corresponding results are presented in Table 3. Since our main concern is to check the moderating effect of the three socio-demographic features, we do not report R-squared values in this table. There are no specific patterns revealed in the significance values of this table. The composite independent variables based on the multiplication of *education* and each independent variable do not show any significant outcome for delta and omicron variants. *Age* moderates the relations the present infection number (*InfNum*_*t*_) has with *RNInf*_*(t-1)*_ and *InfNum*_*(t-1)*_ for only the delta variants. For the omicron variant, *age* moderates only the relation between *InfNum*_*(t-1)*_ and *InfNum*_*t*_. On the other side, *income* moderates the association between *InfNum*_*(t-1)*_ and *InfNum*_*t*_ for both variants.

**Table 3:**
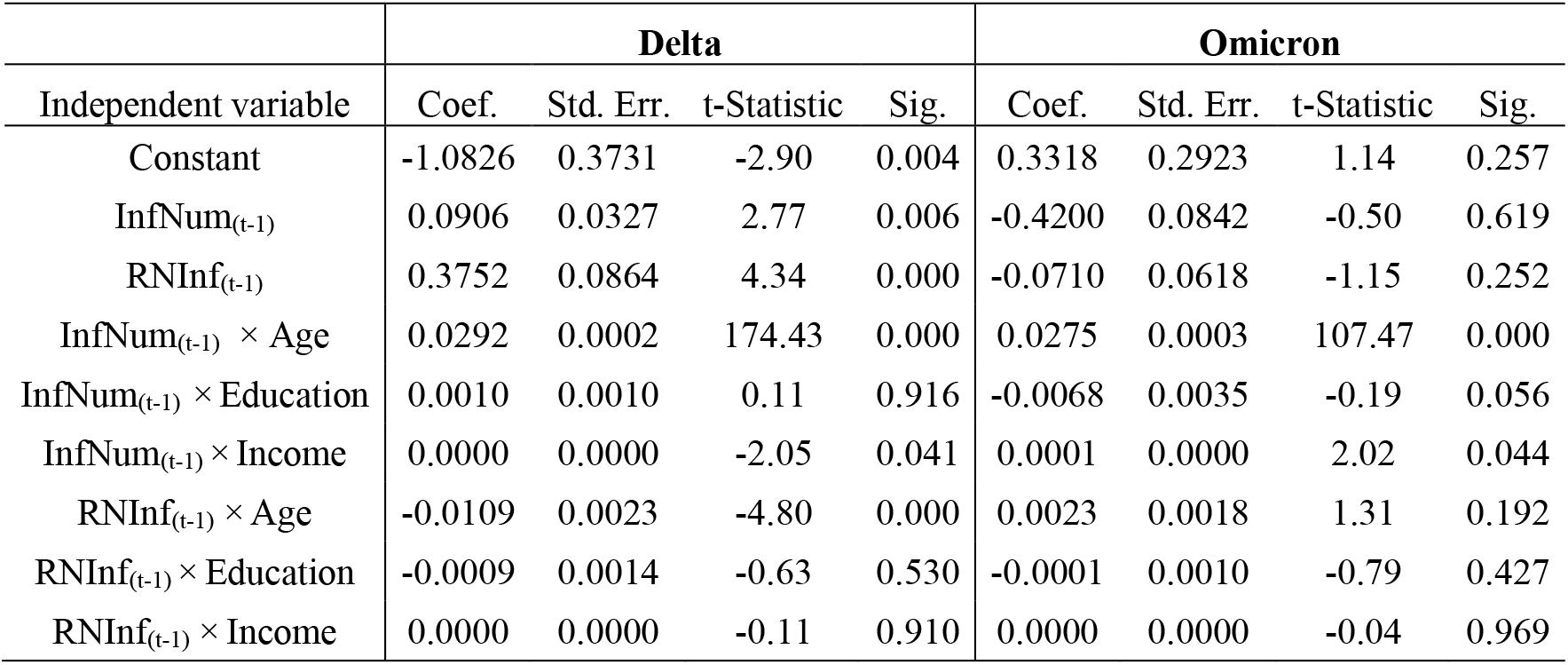
Panel regression outcome for checking the moderating impact of *education* and *income*

| Independent variable | Delta |  |  |  | Omicron |  |  |  |
| --- | --- | --- | --- | --- | --- | --- | --- | --- |
|  | Coef. | Std. Err. | t-Statistic | Sig. | Coef. | Std. Err. | t-Statistic | Sig. |
| Constant | -1.0826 | 0.3731 | -2.90 | 0.004 | 0.3318 | 0.2923 | 1.14 | 0.257 |
| InfNum<sub>(t-1)</sub> | 0.0906 | 0.0327 | 2.77 | 0.006 | -0.4200 | 0.0842 | -0.50 | 0.619 |
| RNInf<sub>(t-1)</sub> | 0.3752 | 0.0864 | 4.34 | 0.000 | -0.0710 | 0.0618 | -1.15 | 0.252 |
| InfNum<sub>(t-1)</sub> × Age | 0.0292 | 0.0002 | 174.43 | 0.000 | 0.0275 | 0.0003 | 107.47 | 0.000 |
| InfNum<sub>(t-1)</sub> × Education | 0.0010 | 0.0010 | 0.11 | 0.916 | -0.0068 | 0.0035 | -0.19 | 0.056 |
| InfNum<sub>(t-1)</sub> × Income | 0.0000 | 0.0000 | -2.05 | 0.041 | 0.0001 | 0.0000 | 2.02 | 0.044 |
| RNInf<sub>(t-1)</sub> × Age | -0.0109 | 0.0023 | -4.80 | 0.000 | 0.0023 | 0.0018 | 1.31 | 0.192 |
| RNInf<sub>(t-1)</sub> × Education | -0.0009 | 0.0014 | -0.63 | 0.530 | -0.0001 | 0.0010 | -0.79 | 0.427 |
| RNInf<sub>(t-1)</sub> × Income | 0.0000 | 0.0000 | -0.11 | 0.910 | 0.0000 | 0.0000 | -0.04 | 0.969 |

Figure 3 shows the kernel density estimation (KDE) for *age, education* and *income*. KDE is a non-parametric way to estimate the probability density function of a random variable (Terrell and Scott 1992). The median value of each socio-demographic attribute is used to split the dataset into two groups. The density estimations are based on this study’s single dependent variable (*InfNum*_*t*_), divided into two groups by each of the three socio-demographic attributes. This figure reveals that the density functions are closely identical between different groups based on *age, education* and *income*, which further echos the findings from Table 3. These three socio-demographic attributes do not reveal any specific patterns in moderating the relationship between the model’s independent and dependent variables.

**Figure 3:**
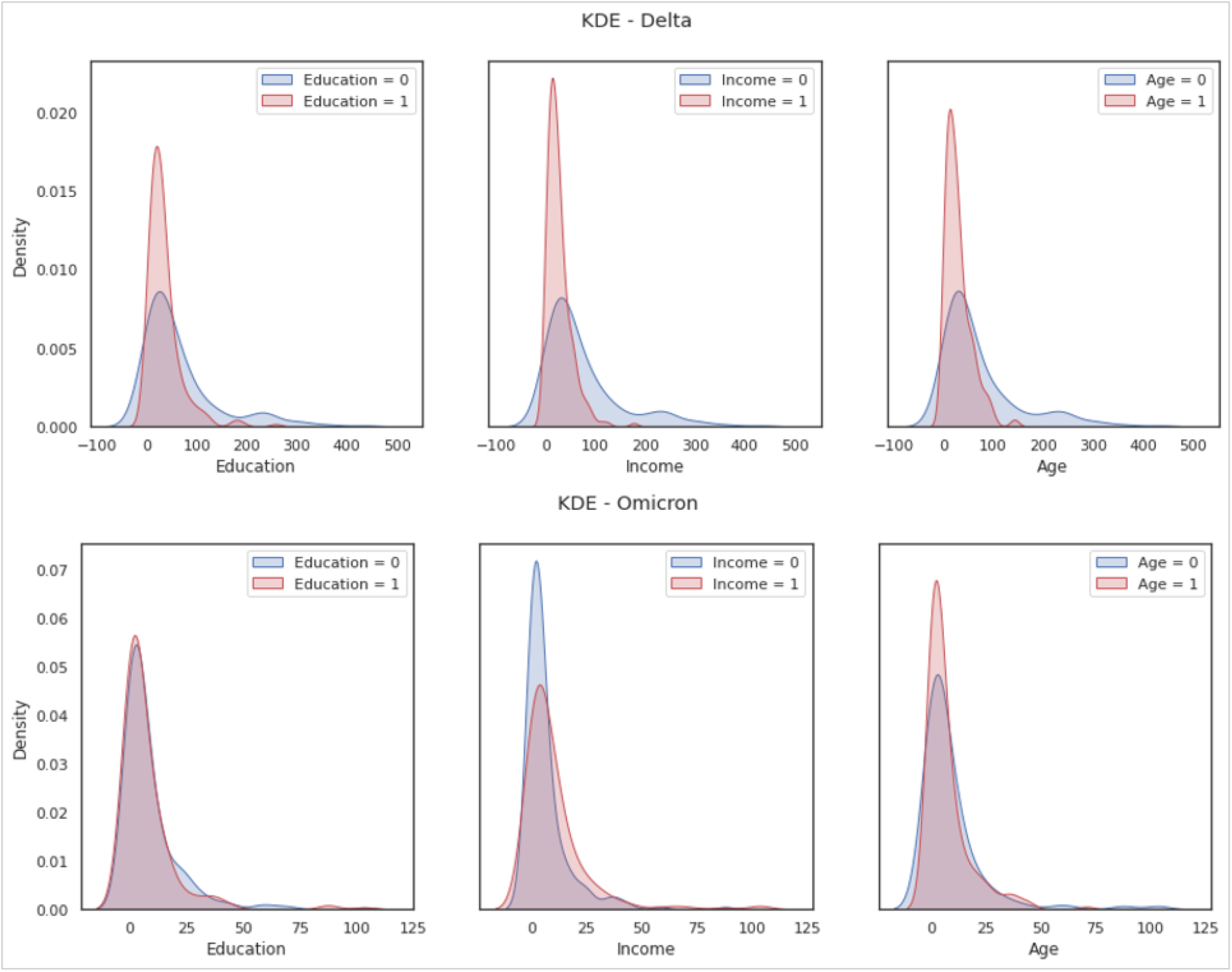
The kernel density estimation of the independent variable (*InfNumt*) based on the socio-demographic attributes of *age, education* and *income*

## 5.Discussion

Human mobility data has been shown to be an effective measure for modelling COVID-19 infection count (Hou et al., 2021). In the first part of this study, we aimed to capture this mobility through the neighbourhood measure and its effect on COVID-19 infection count. The *neighbourhood measure* considered a relatively granular suburb level as a geographical unit and used the number of shared roads to approximate human movement across the suburbs. The research dataset covers two periods of COVID-19 infection for delta and omicron variants, as shown in Table 1 earlier. One interesting perspective to note and explore in this study is that some of the underlying factors changed between these two timeframes. During the delta outbreak, the research areas were under lockdown (with only allowed shopping limit within a 5 km radius for essential items). Some areas of concern even had nighttime curfew during this timeframe. Sydney’s vaccination coverage (double dose) went from approximately 26% to 43% (Australian Broadcasting Corporation News 2022). On the other hand, there was no lockdown during the omicron phase of the dataset, although mask mandates, social distancing, and capacity caps in businesses partially remained (Reuters News 2022). Double dose vaccination coverage (double dose) rose from 77% to almost 79% during this period. As a result, it was inevitable that people’s mobility within and across the suburbs during the Omicron outbreak was significantly higher. The omicron variant itself is more transmissible than the delta variant. Therefore, it would be interesting to see how the neighbourhood measure affected the infection count during delta and omicron outbreaks.

The fixed effect panel regression model shows good prediction performance for the delta variant with an R-squared value of 85.66%. The model performance was relatively weaker for the omicron variant with a 52.67% R-squared value. The previous infection count has a significant positive impact on the present infection count (dependent variable) for both variants. The same goes for the neighbourhood measure on its impact on present infection count except that for delta, the effect is positive, and for omicron, it is negative. Together these results indicate that infection count for a suburb during the delta variant can be well modelled through past infection count and influx from surrounding suburbs, i.e., *neighbourhood measure*. While present infection count should naturally be affected by previous infection count, the impact of influx from the neighbourhood is more interesting. As we mentioned earlier, especially during the delta outbreak, there was a lockdown in place, and residents were only allowed to go out for essential shopping within a 5km radius. Suburbs in our research are relatively granular in size, and residents could move across the neighbouring suburbs for essential reasons even with staying within a 5km bubble. Therefore, this prediction model using suburb-level granular data effectively captures macro-movement during the lockdown and utilizes it to predict case count during delta variant.

For the omicron variant, the regression model and the neighbourhood measure did not reveal many insights because the R-square value was not much higher than the delta variant and the neighbourhood measure showed a significant negative impact on infection count counter-intuitively. Two factors could contribute to this finding. First, there was no lockdown or movement restriction during the omicron variant. Second, omicron is more transmissible compared to the delta variant (Cameroni et al., 2021). The high contagiousness and unrestricted movement within the suburb might make the neighbourhood measure less reliable in predicting the case count for omicron.

In the second part of this research, we looked into three socio-economic moderating factors - age, education and income. We intended to see whether suburbs with more residents of higher age bracket, education level or income differ from suburbs having fewer residents with those factors in terms of case count and neighbourhood measure. This was important in a way that during the delta outbreak, a lockdown was imposed in the areas of concern and a nighttime curfew for some period of time. These areas of concern were mostly concentrated in western Sydney, where a large proportion of the residents are culturally and linguistically diverse and have a migrant background. These suburbs have more members per household, less income, and education level on average. Many of the wage earners’ jobs could not be performed from home. Consequently, stay-at-home orders and the lockdown hard hit these suburbs more (Australian Broadcasting Corporation News 2022). Therefore, we investigated these suburbs with high population and COVID-19 cases and explored whether age, education and income have any moderating effect on the case count and neighbourhood measure.

The results in the earlier Table 3 summarily shows the moderating effects. Education did not have any moderating effect for any combination. For both delta and omicron variants, age and income both had significantly moderated the relation between previous and present case counts. However, income has a small coefficient value for the moderating effect and thus does not reveal any meaningful insight. Age has a positive coefficient indicating that suburbs having a population of higher age bracket tended to have higher case growth. This goes along with the fact that older people are at higher risk of comorbidities and COVID-19 (Monod et al., 2021). Age positively moderates the relation between neighbourhood measure and present case count only for the delta variant. This might indicate that suburbs with a relatively higher aged population tend to have more mobility (for work or essential purposes) if they have more options to travel across suburbs through the higher number of available road connections. For the omicron variant, we have seen earlier that the neighbourhood measure does not affect the case count, probably due to the high transmissibility of the variant and significant local movement due to the absence of lockdown. Consequently, none of the socio-economic variables moderated the relation between the neighbourhood measure and case count.

## 6. Conclusion

The Greater Sydney area residents endured nearly four months of COVID-19 lockdown during the last half of 2021. While the lockdown bought precious time to ramp up vaccination rollout and prepare healthcare facilities, it left a lasting economic and psychological impact. This study analyzed the mobility and prevalence data in two distinct timeframes to model and predicted the COVID-19 case count during late 2021. The timeframes represented delta and omicron outbreaks, respectively, and for the former outbreak, there was lockdown in place and nighttime curfew for some period. The road network between the neighbouring suburbs was used to approximate the influx and corresponding risk of case growth from adjacent areas. Therefore, this study helps us to explore and compare the effect of mobility and case count during a lockdown and without lockdown period. It also gives a comparison between delta and omicron variants. The moderating effect of three socio-economic variables is discussed. The methods introduced in this study shows an effective way to utilize geographic information and road connection network with health data to model COVID-19 transmission. The regression model results show that the road network-based neighbourhood measure significantly predicts the case count for the delta variant. The results also show that the income or education level of the residents do not necessarily have any effect in moderating the case count and mobility. The methodology presented in this study could be replicated for other states or countries to gather similar insights.

## Data Availability

Data are taken from publicly available sources

## Notes

### Competing Interest Statement

The authors have declared no competing interest.

### Funding Statement

This study did not receive any funding

## References

Abdy, M., Side, S., Annas, S., Nur, W. and Sanusi, W. (2021). “An SIR epidemic model for COVID-19 spread with fuzzy parameter: the case of Indonesia.” Advances in Difference Equations 2021(1).

Al Wahaibi, A., et al. (2021). “The impact of mobility restriction strategies in the control of the COVID-19 pandemic: modelling the relation between COVID-19 health and community mobility data.” International Journal of Environmental Research and Public Health 18(19): 10560.

Australian Broadcasting Corporation News (2022). “How Sydney’s COVID-19 lockdown is dividing the city.” from https://www.abc.net.au/news/2021-08-22/sydney-covid-19-lockdown-is-creating-growing-inequality/100391922.

Australian Broadcasting Corporation News (2022). “Tracking Autralia’s COVID vaccine rollout numbers.” Retrieved February 26, 2022, from https://www.abc.net.au/news/2021-03-02/charting-australias-covid-vaccine-rollout/13197518.

Cameroni, E., et al. (2021). “Broadly neutralizing antibodies overcome SARS-CoV-2 Omicron antigenic shift.” Nature: 1–9.

Car, Z., Baressi Šegota, S., Andelic, N., Lorencin, I. and Mrzljak, V. (2020). “Modeling the Spread of COVID-19 Infection Using a Multilayer Perceptron.” Computational and Mathematical Methods in Medicine 2020: 5714714.

Census QuickStats (2021). “Australian Bureau of Statistics: 2016 Census QuickStats.” Retrieved May 25, 2021, from https://quickstats.censusdata.abs.gov.au/census_services/getproduct/census/2016/quickstat/POA2190?opendocument.

Chimmula, V. K. R. and Zhang, L. (2020). “Time series forecasting of COVID-19 transmission in Canada using LSTM networks.” Chaos, solitons, and fractals 135: 109864–109864.

Google maps (2021). “Google maps, Australia.” Retrieved June 15, 2021, from www.maps.google.com.au.

Hernandez-Matamoros, A., Fujita, H., Hayashi, T. and Perez-Meana, H. (2020). “Forecasting of COVID19 per regions using ARIMA models and polynomial functions.” Applied Soft Computing 96: 106610.

Hou, X., Gao, S., Li, Q., Kang, Y., Chen, N., Chen, K., Rao, J., Ellenberg, J. S. and Patz, J. A. (2021). “Intracounty modeling of COVID-19 infection with human mobility: Assessing spatial heterogeneity with business traffic, age, and race.” 118(24): e2020524118.

Kohler, U. and Kreuter, F. (2005). Data analysis using Stata, Stata press.

Liu, X. X., Fong, S. J., Dey, N., Crespo, R. G. and Herrera-Viedma, E. (2021). “A new SEAIRD pandemic prediction model with clinical and epidemiological data analysis on COVID-19 outbreak.” Applied Intelligence 51(7): 4162–4198.

Lotfi, M., Hamblin, M. R. and Rezaei, N. (2020). “COVID-19: Transmission, prevention, and potential therapeutic opportunities.” Clinica chimica acta 508: 254–266.

Melin, P., Monica, J. C., Sanchez, D. and Castillo, O. (2020). “Multiple Ensemble Neural Network Models with Fuzzy Response Aggregation for Predicting COVID-19 Time Series: The Case of Mexico.” Healthcare 8(2).

Monod, M., et al. (2021). “Age groups that sustain resurging COVID-19 epidemics in the United States.” science 371(6536): eabe8372.

Nižetic, S. (2020). “Impact of coronavirus (COVID-19) pandemic on air transport mobility, energy, and environment: A case study.” International Journal of Energy Research 44(13): 10953–10961.

NSW Health (2021). “COVID-19 data and statistics.” Retrieved December 25, 2021, from https://www.nsw.gov.au/covid-19/stay-safe/data-and-statistics.

Pinter, G., Felde, I., Mosavi, A., Ghamisi, P. and Gloaguen, R. (2020). “COVID-19 Pandemic Prediction for Hungary; A Hybrid Machine Learning Approach.” Mathematics 8(6).

Reuters News (2022). “Freedom Day’: Sydney reopens as Australia looks to live with COVID-19.” Retrieved February 26, 2022, from https://www.reuters.com/world/asia-pacific/long-100-days-sydney-reopens-australia-looks-live-with-covid-19-2021-10-10/.

Štifanić, D., Musulin, J., Miočević, A., Baressi Šegota, S., Šubić, R. and Car, Z. (2020). “Impact of COVID-19 on Forecasting Stock Prices: An Integration of Stationary Wavelet Transform and Bidirectional Long Short-Term Memory.” Complexity 2020: 1846926.

Sumner, A., Hoy, C. and Ortiz-Juarez, E. (2020). Estimates of the Impact of COVID-19 on Global Poverty, WIDER working paper.

Swaraj, A., Verma, K., Kaur, A., Singh, G., Kumar, A. and Melo de Sales, L. (2021). “Implementation of stacking based ARIMA model for prediction of Covid-19 cases in India.” J Biomed Inform 121: 103887.

Terrell, G. R. and Scott, D. W. (1992). “Variable kernel density estimation.” The Annals of Statistics 20(3): 1236–1265.

Yudistira, N. (2020). “COVID-19 growth prediction using multivariate long short term memory.” arXiv preprint arXiv:2005.04809.

Zhou, Y., Xu, R., Hu, D., Yue, Y., Li, Q. and Xia, J. (2020). “Effects of human mobility restrictions on the spread of COVID-19 in Shenzhen, China: a modelling study using mobile phone data.” The Lancet Digital Health 2(8): e417–e424.

